# Observational Study of Metformin and Risk of Mortality in Patients Hospitalized with Covid-19

**DOI:** 10.1101/2020.06.19.20135095

**Authors:** Carolyn T. Bramante, Nicholas E. Ingraham, Thomas A. Murray, Schelomo Marmor, Shane Hovertsen, Jessica Gronski, Chace McNeil, Ruoying Feng, Gabriel Guzman, Nermine Abdelwahab, Samantha King, Thomas Meehan, Kathryn M. Pendleton, Bradley Benson, Deneen Vojta, Christopher J. Tignanelli

**Affiliations:** Department of Medicine, University of Minnesota, Division of General Internal Medicine, Minneapolis, MN; Department of Medicine, University of Minnesota, Division of Pulmonary, Allergy, Critical Care and Sleep Medicine, Minneapolis, MN; School of Public Health, University of Minnesota, Division of Biostatistics, Minneapolis, MN; Department of Surgery, University of Minnesota Division of Surgical Oncology, Minneapolis, MN; Department of Surgery, University of Minnesota Division of Acute Care Surgery, Minneapolis, MN; Research and Development, UnitedHealth Group; Institute for Health Informatics, University of Minnesota, Minneapolis, MN

**Author notes:** Authors contributed equally to this manuscript (Carolyn T. Bramante, Nicholas E. Ingraham, Deneen Vojta, Christopher J. Tignanelli). Correspondence: Carolyn T. Bramante MD, MPH, 420 Delaware St SE, MMC 276, Minneapolis, MN 55455, Office: (612).

## Abstract

**Importance:** Type 2 diabetes (T2DM) and obesity are significant risk factors for mortality in Covid19. Metformin has sex specific immunomodulatory effects which may elucidate treatment mechanisms in COVID-19. **Objective:** We sought to identify whether metformin reduced mortality from Covid19 and if sex specific interactions exist.

**Design:** Retrospective review of de-identified claims from UnitedHealth Group’s Clinical Discovery Database. Unadjusted and multivariate models were conducted to assess risk of mortality based on metformin and tumor necrosis factor alpha (TNFα) inhibitors as home medications in individuals with T2DM and obesity, controlling for comorbidities, medications, demographics, and state. Heterogeneity of effect was assessed by sex.

**Setting:** The database includes all 50 states in the United States. **Participants:** Persons with at least 6 months of continuous coverage from UnitedHealth Group in 2019 who were hospitalized with Covid-19. Persons in the metformin group had > 90 days of metformin claims in the 12 months before hospitalization.

**Results:** 6,256 persons were included; 52.8% female; mean age 75 years. Metformin was associated with decreased mortality in women by logistic regression, OR 0.792 (0.640, 0.979); mixed effects OR 0.780 (0.631, 0.965); Cox proportional-hazards: HR 0.785 (0.650, 0.951); and propensity matching, OR of 0.759 (0.601, 0.960). There was no significant reduction in mortality among men. TNFα inhibitors were associated with decreased mortality, by propensity matching in a limited model, OR 0.19 (0.0378, 0.983).

**Conclusions:** Metformin was significantly associated with reduced mortality in women with obesity or T2DM in observational analyses of claims data from individuals hospitalized with Covid-19. This sex-specific finding is consistent with metformin’s reduction of TNFα in females over males, and suggests that metformin conveys protection in Covid-19 through TNFα effects. Prospective studies are needed to understand mechanism and causality.

**Key Points:** *Question:* Metformin has many anti-inflammatory effects, including sex-specific effects on TNFα. Is metformin protective from the Sars-CoV-2 virus, and does the effect differ by sex?

*Findings:* Metformin was associated with reduced mortality in women who were hospitalized with Covid-19, but not in men who were hospitalized with Covid-19.

*Meaning:* The sex-dependent survival by metformin use points towards TNFα reduction as a key mechanism for protection from Covid-19.

## Introduction

The coronavirus disease 2019 (Covid-19), caused by the severe acute respiratory syndrome coronavirus-2 (SARS-CoV-2), has spread throughout the world.^1^ Despite exponential growth in Covid-19 related research, better understanding of this highly contagious and lethal virus is needed. An overall mortality rate over 5% for all patients hospitalized with Covid-19 highlights the urgent need for treatments while vaccines are developed.^2^

Observational data early in the outbreak identified male sex and increased age as leading risk factors in Covid-19.^3^ Subsequent studies have identified hypertension, diabetes, coronary artery disease, tobacco use, and obesity as important risk factors for Covid-19 outcomes.^2,4-6^ Persons hospitalized with Covid-19 with overweight or obesity (body mass index, BMI≥25kg/m^2^) had a higher risk of needing mechanical ventilation, after controlling for diabetes, hypertension, and cardiovascular disease.^6^ Additionally, among individuals with Covid-19 and a BMI ≥28kg/m,^2^ men have a higher risk of developing severe Covid-19 than women.^7^ This sex difference may be explained by the accumulation of visceral adiposity at lower BMI levels in men.8

Adipocytes, specifically visceral adipocytes, secrete many of the inflammatory and coagulopathic molecules that are implicated in Covid-19 morbidity, including interleukin-6 (IL-6), tumor-necrosis-factorα (TNFα), d-dimer, and others.^9-12^ TNFα has been particularly important, with high levels of TNFα found in lung tissue of persons with Covid-19.^13^ TNFα contributes to insulin resistance, and levels of TNFα are higher in individuals with type 2 diabetes (T2DM).^14^ Both T2DM and obesity are associated with lower levels of the anti-inflammatory cytokine, IL-10.^15^ Metformin, the first-line medication for type 2 diabetes (T2DM) decreases TNFα and IL-6, boost levels of IL-10, and has been found to cause these beneficial effects significantly more in females more than males.^16-20^ Metformin also increases activation of AMP-activated protein kinase (AMPK), which has important downstream effects in Covid-19.^16,21,22^

Given these favorable effects of metformin on TNFα and other inflammatory cytokines that contribute to Covid-19, our primary objective was to understand whether home metformin use was associated with decreased mortality in persons hospitalized with Covid-19. We hypothesized that metformin would be associated with decreased mortality from Covid-19 in persons with T2DM or obesity, and that this benefit would be higher in women compared to men given metformin’s sex-specific anti-inflammatory effects. We also hypothesized that TNFα inhibitors would be associated with decreased mortality from Covid-19. We conducted a retrospective cohort analysis of de-identified claims data from UnitedHealth Group’s Clinical Discovery Database of 6,256 persons hospitalized in the US with Covid-19 in 2020.

## Methods

### Design and Data

Retrospective analysis of claims from UnitedHealth Group (UHG)’s Clinical Discovery Database between January 1, 2020 – June 7, 2020. This database includes de-identified individual-level and state-level data for individuals with Covid-19 admissions in all 50 U.S. states, covering a diverse range of ages, ethnicities, and geographical regions. The claims data includes medical and pharmacy claims, laboratory results, and enrollment records. This study was approved by the University of Minnesota institutional review board (STUDY00001489) which provided a waiver of consent for this study.

### Population

Individuals 18 years or older with T2DM or obesity, at least 6 months of continuous enrollment in 2019, and a hospitalization for Covid-19 confirmed by polymerase change reaction (PCR), manual chart review by UHG, or reported from the hospital to UHG. Individuals with both commercial and Medicare Advantage insurance were included. Eighteen persons (0.12%) were missing age and were excluded. An assumption was made that there was no missingness among the other variables. Individuals with T2DM or obesity were included in the analysis (definition in Supplemental Materials 1).

### Outcomes

Our primary outcome was in-hospital mortality defined using the hospital disposition indicator. The individuals who remained hospitalized on June 7, 2020 without a hospital disposition were censored in Cox proportional hazards model analyses and excluded from mixed model and propensity-matched analyses. The database did not include data related to in-hospital complications, ICU, or ventilator utilization and thus an analysis on these endpoints was not possible.

### Independent Variable and confounding variables

The independent variable was a filled pharmacy prescription (visible through prescription claims, matching the generic drug ingredient with the string “metformin”) indicating use of metformin, and limited to patients with at least 90 days of metformin prescription from 12 months prior to Covid-19 diagnosis. Potential confounding medications hypothesized to be protective or harmful for patients with Covid-19 determined by a large evidence-based consortium were assessed, as were possible confounding co-morbidities (Supplemental Materials 1).^23^

### Statistical analysis

Cohort age was expressed by median and interquartile range (IQR). Categorical variables were expressed by percentages. Univariate analysis compared mortality for persons without vs with home metformin use.

To determine whether metformin use was independently associated with reduced mortality for patients hospitalized for Covid-19, multivariable models for mortality were developed with the use of the Least absolute shrinkage and selection operator (LASSO) method,^24^ with the tuning parameter determined by the Akaike information criterion (AIC). We also considered clinically relevant pairwise interactions to determine whether their association with mortality differed according to metformin status. The non-linear effect of age was modeled in two manners, using restricted cubic splines or categorized as (0-55, 56-65, 66-75, 76-85, 86+). Given the low mortality for patients 0-55 (5.1%) compared with other age groupings, this group was not further categorized. Subgroup analyses of multivariate models were performed by sex.

### Multivariate models

1. Logistic regression, controlling for all covariates in our conceptual framework (Table 1), and for LASSO variables and state, with and without specific disease-medication interaction terms. (eTable 1).
2. Mixed-effects logistic regression with state-level random effects, controlling for LASSO variables, with and without specific disease-medication interaction terms. Only patients with known hospital disposition were included in this analysis (eTable 2).
3. Cox proportional-hazards regression by strata-specific and shared-frailty effects, with and without specific disease-medication interaction terms; censoring determined based on claims made after hospitalization up to June 7, 2020. Using a ‘best’ outcome approach, patients discharged not to hospice were assigned a censoring time equal to the longest observed hospital stay (169 days).^25,26^ Scaled Schoenfeld residual graphs and log-log plots were analyzed to confirm adequacy of the proportional hazards assumption for home metformin use (eTable 3, eFigure 1).
4. Propensity matched mixed effects logistic regression was performed, stratified by metformin use. Propensity scores were estimated with logistic regression with variables selected by the aforementioned LASSO logistic model and two evenly matched groups were formed with the common caliper set at 0.2 (eFigure 2), and a model with exact matching.^27,28^ Even distribution of propensity scores was confirmed between matched groups, with standardized differences less than 0.1 for all confounding variables (eFigure 3). Univariate logistic regression was then used to compare mortality for persons who were receiving (vs not receiving) home metformin among the matched cohort. Kaplan-Meier survival curves were also estimated and compared using a log-rank test (Figure 2).

**Table 1:** Demographic and clinical characteristics of patients hospitalized for Covid-19 with 6 months of continuous insurance coverage in 2019, comparing those on home metformin to those not on metformin.

| **Demographic characteristics** | **Metformin** |  |
| --- | --- | --- |
|  | **No (n=3,923)** | **Yes (n=2,333)** |
| Age, median (inter-quartile range) | 76.0 (67.0, 84.0) | 73.0 (66.0, 80.0) |
| Age under 56 years, n (%) | 266 (6.8%) | 186 (8.0%) |
| Age 56-65 years | 535 (13.6%) | 387 (16.6%) |
| Age 66-75 years | 1112 (28.3%) | 808 (34.6%) |
| Age 76-85 years | 1234 (31.5%) | 694 (29.7%) |
| Age > 85 years | 776 (19.8%) | 258 (11.1%) |
| Female sex, n (%) | 2,173 (55.4%) | 1,129 (48.4%) |
| Transfer,<sup>b</sup> n (%) | 705 (18.0%) | 418 (17.9%) |
| **Pre-existing Conditions, n (%)** |  |  |
| Type 2 Diabetes | 3719 (94.8%) | 2316 (99.3%) |
| Type 1 Diabetes | 278 (7.1%) | 108 (4.6%) |
| Essential hypertension | 2370 (60.4%) | 1314 (56.3%) |
| Tobacco | 8 (0.2%) | 5 (0.2%) |
| Coronary artery disease | 864 (22.0%) | 456 (19.5%) |
| Heart failure with preserved ejection fraction | 346 (8.8%) | 121 (5.2%) |
| Heart failure with reduced ejection fraction | 344 (8.8%) | 133 (5.7%) |
| Heart failure, unspecified | 659 (16.8%) | 239 (10.2%) |
| Liver disease | 160 (4.1%) | 102 (4.4%) |
| Venous thromboembolism | 161 (4.1%) | 62 (2.7%) |
| Neutropenia | 9 (0.2%) | 5 (0.2%) |
| Cancer | 441 (11.2%) | 281 (12.0%) |
| Coagulation defect | 54 (1.4%) | 14 (0.6%) |
| Valve Repair | 33 (0.8%) | 17 (0.7%) |
| Chronic obstructive pulmonary disease | 688 (17.5%) | 305 (13.1%) |
| Interstitial lung disease | 70 (1.8%) | 33 (1.4%) |
| Chronic kidney disease, stage 3, 4 | 729 (18.6%) | 147 (6.3%) |
| Chronic kidney disease, unspecified | 527 (13.4%) | 219 (9.4%) |
| End stage renal disease | 297 (7.6%) | 14 (0.6%) |
| Atrial fibrillation | 630 (16.1%) | 290 (12.4%) |
| Cerebrovascular accident/transient ischemic attack | 432 (11.0%) | 207 (8.9%) |
| Alcohol abuse | 43 (1.1%) | 17 (0.7%) |
| Human immunodeficiency virus (HIV) | 18 (0.5%) | 18 (0.8%) |
| Asthma | 166 (4.2%) | 96 (4.1%) |
| General influenza | 65 (1.7%) | 35 (1.5%) |
| Swine, avian influenza | 17 (0.4%) | 12 (0.5%) |
| Inflammatory bowel disease | 28 (0.7%) | 11 (0.5%) |
| Systemic lupus erythematosus, rheumatoid arthritis | 73 (1.9%) | 27 (1.2%) |
| Dementia | 663 (16.9%) | 273 (11.7%) |
| Charlson comorbidity index, median score (IQR) | 5.0 (3.0, 7.0) | 4.0 (3.0, 6.0) |
| Diabetes complications severity index, median (IQR) | 2.0 (1.0, 4.0) | 2.0 (0.0, 3.0) |
| Death | 791 (21.3%) | 394 (17.8%) |
| **Body Mass Index Category** |  |  |
| Absence of any weight-related code | 3548 (90.4%) | 2215 (94.9%) |
| Overweight (BMI 25-30) | 21 (0.5%) | 7 (0.3%) |
| Class I obesity (BMI 31-35) | 35 (0.9%) | 10 (0.4%) |
| Class II obesity (BMI 36-40) | 20 (0.5%) | 6 (0.3%) |
| Class III obesity (BMI 40+) | 190 (4.8%) | 62 (2.7%) |
| Obesity unspecified | 109 (2.8%) | 33 (1.4%) |
| **Medications** |  |  |
| Ursodiol | 8 (0.2%) | 2 (0.1%) |
| Angiotensin-converting enzyme inhibitors | 1069 (27.2%) | 912 (39.1%) |
| Angiotensin II receptor blocker | 1003 (25.6%) | 731 (31.3%) |
| Statin | 2591 (66.0%) | 1860 (79.7%) |
| Antiplatelet | 618 (15.8%) | 342 (14.7%) |
| Anticoagulation | 808 (20.6%) | 407 (17.4%) |
| Tenofovir | 5 (0.1%) | 5 (0.2%) |
| Highly Active Antiretroviral Therapy (HAART) | 16 (0.4%) | 14 (0.6%) |
| Azithromycin | 541 (13.8%) | 321 (13.8%) |
| Second line diabetes medications | 27 (0.7%) | 35 (1.5%) |
| Insulin | 1564 (39.9%) | 783 (33.6%) |
| Steroids | 1010 (25.7%) | 546 (23.4%) |
| Hydroxychloroquine | 47 (1.2%) | 14 (0.6%) |
| Janus Kinase Inhibitors | 3 (0.1%) | 1 (<1%) |
| Calcineurin Inhibitors | 85 (2.2%) | 31 (1.3%) |
| mTor Inhibitor | 1 (<1%) | 0 (0.0%) |
| Beta Blocker | 2136 (54.4%) | 1213 (52.0%) |
| Ivermectin | 35 (0.9%) | 13 (0.6%) |
| Beta2 Agonist | 1233 (31.4%) | 608 (26.1%) |
| Allopurinol | 400 (10.2%) | 189 (8.1%) |
| Azathioprine & Mycophenolate mofetil | 42 (1.1%) | 13 (0.6%) |
| Montelukast | 289 (7.4%) | 181 (7.8%) |
| Nonsteroidal anti-inflammatory drugs | 362 (9.2%) | 316 (13.5%) |
| Diuretics | 1604 (40.9%) | 670 (28.7%) |
| Mast cell stabilizer | 65 (1.7%) | 41 (1.8%) |
| Valacyclovir, acyclovir, valgancyclovir | 129 (3.3%) | 72 (3.1%) |

<sup>a</sup> Transfer represents inter-hospital transfer during the hospitalization for COVID-19.

**Table 2.**
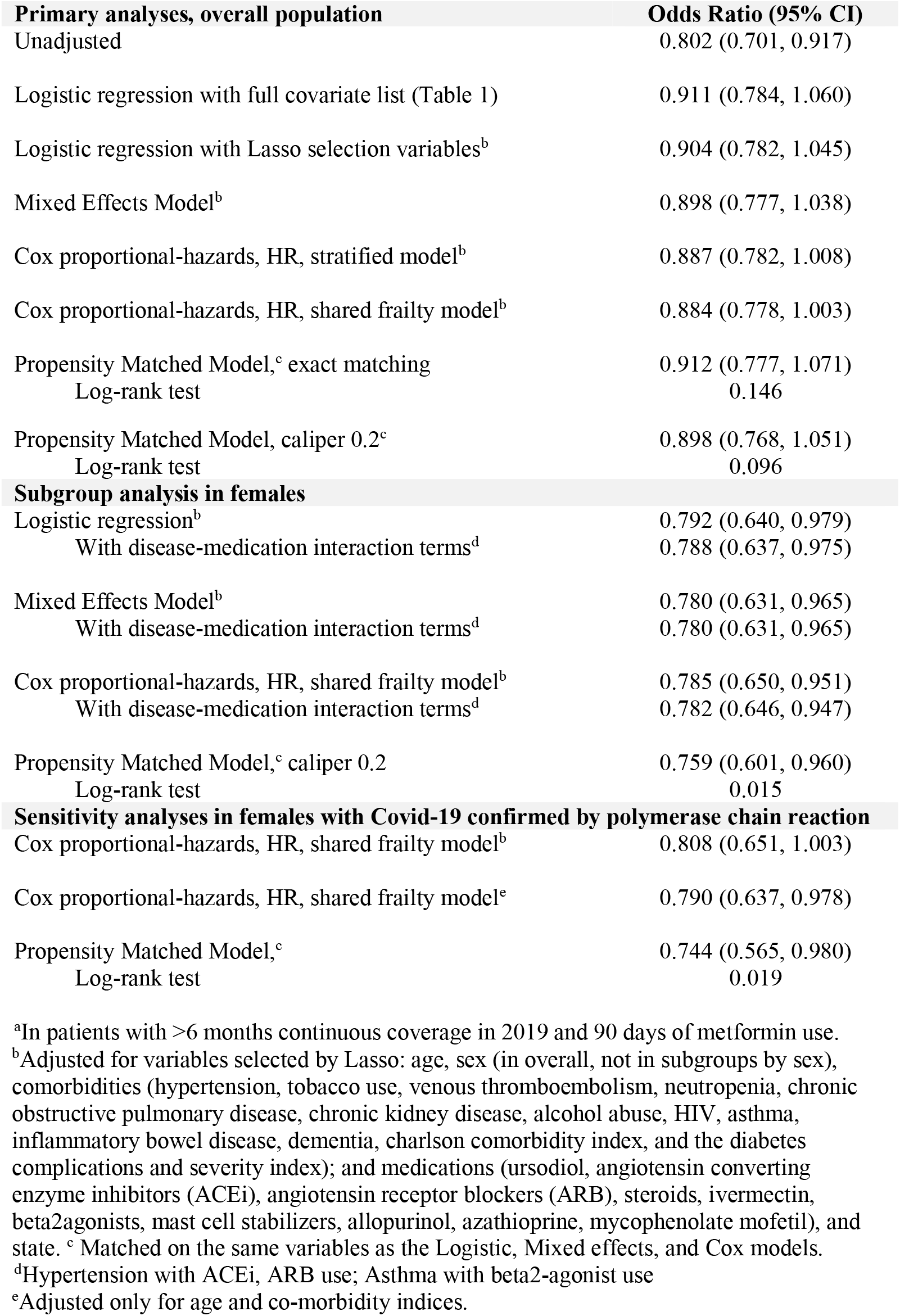
Association between home metformin use and mortality in unadjusted and adjusted analyses in patients with type 2 diabetes or obesity, hospitalized for Covid-19 (confirmed or presumed).^a^

**Figure 1:**
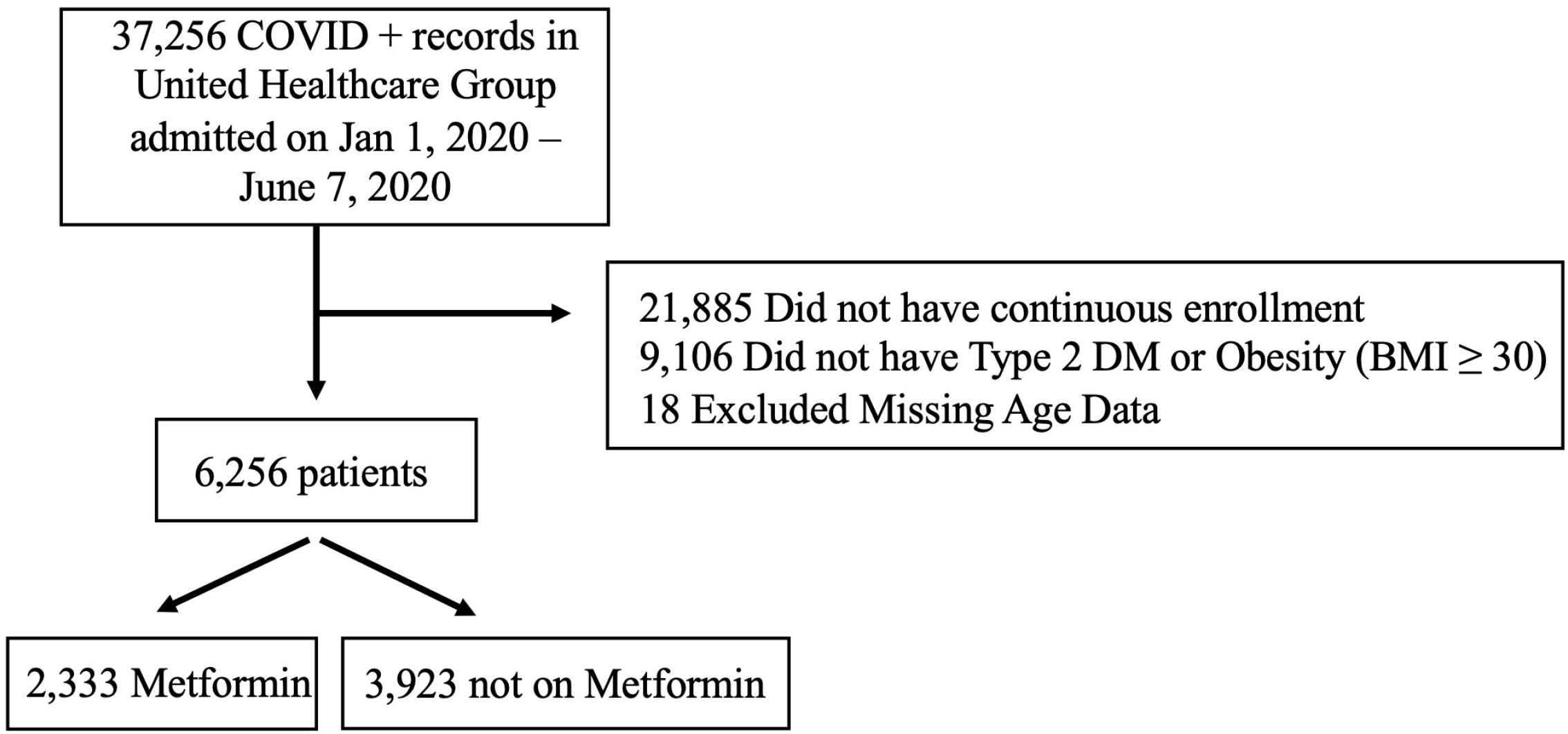
Study diagram detailing selection of patients in United Healthcare Covid-19 Database. This is a flow diagram representing how patients were selected into the analysis.

**Figure 2.**
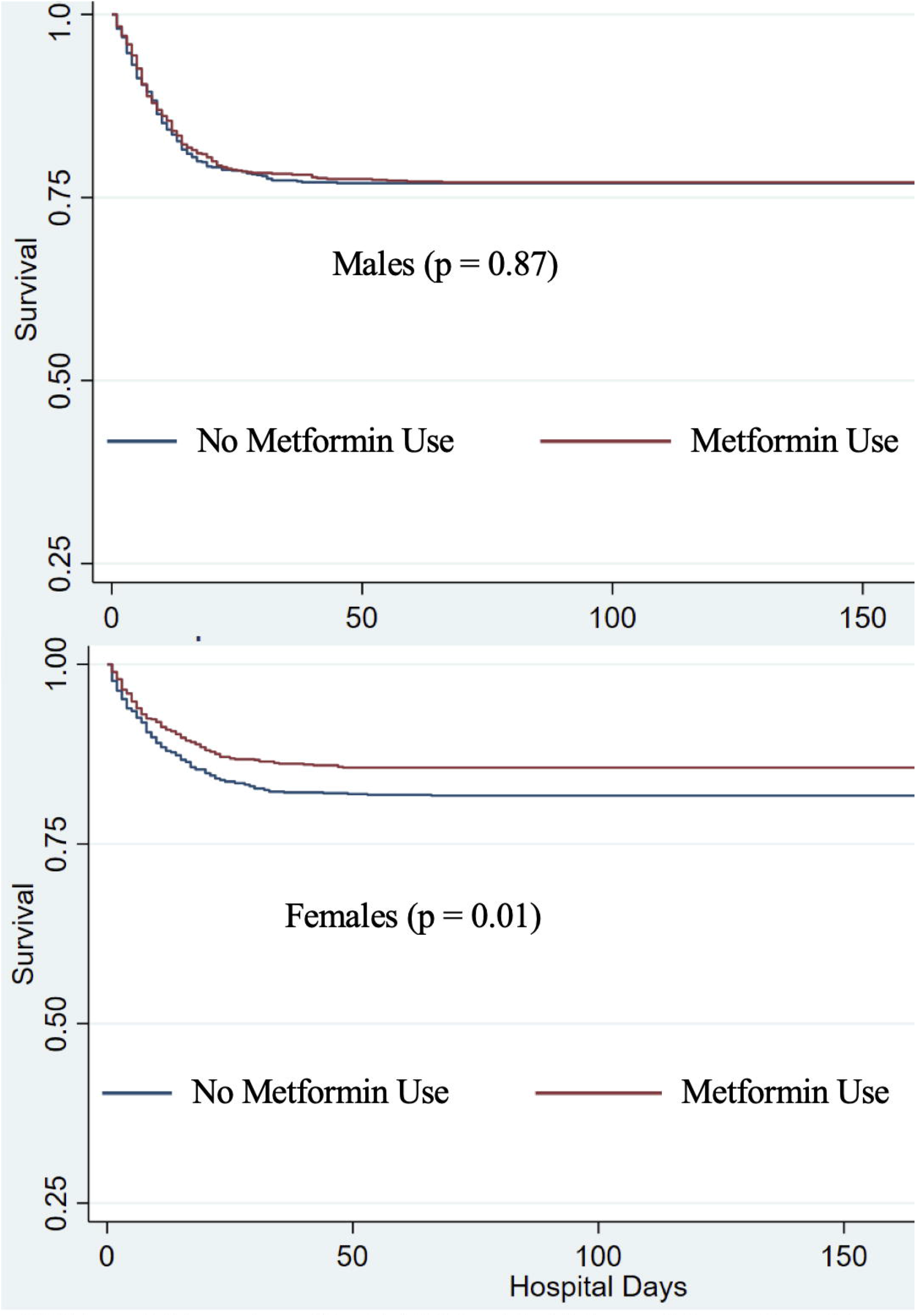
Kaplan Meier curve by metformin use, with propensity matching in persons with type 2 diabetes and obesity hospitalized for Covid-19. *. *With variables selected by LASSO with AlC, matching caliper 0.2 This is a Kaplan-Meier survival curve. The top panel compares metformin use to no metformin use in males only. The bottom panel compares metformin use to no metformin use in females onluy. In both panels, the blue line represents no metformin use and the red line represent metformin use. The Y axis is survival, the X axis is hospital days.

**Figure 3.**
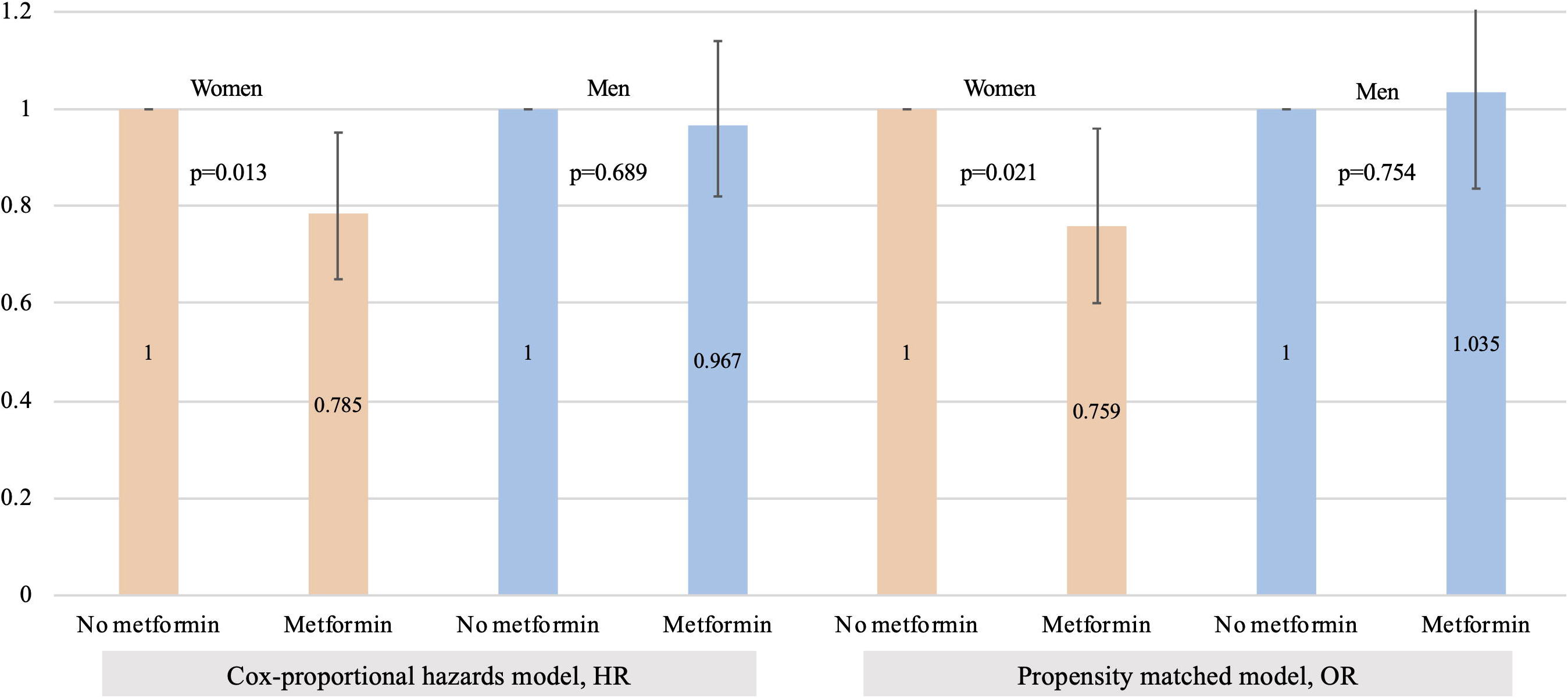
Subgroup analysis of mortality associated with metformin use vs no metformin use in hospitalizations for Covid-19, among women and men with type 2 diabetes or obesity. Bars represent 95% Cl’s. Bar graph comparing survival among women and among men, comparing those without metformin to those with metformin. The left two columns are women, without and with metformin, analyzed by cox proportional hazards. The 3^rd^ and 4^th^ columns (blue) are men, without and with metformin, analyzed by Cox proportional-hazards. The 5^th^ and 6^th^ columns (light orange), are women, without and with metformin, analyzed by propensity matching. The 7^th^ and 8^th^ columns (blue) are men, without and with metformin, analyzed by propensity matching.

Sensitivity analyses were conducted in individuals with Covid-19 confirmed by PCR (n=4,105). Statistical analyses were performed using Stata MP, version 16 (StataCorp, College Station, TX). Statistical significance was defined as a two-tailed *p*-value < 0.05.

## Results

### Characteristics of Cohort

Between Jan 1, 2020 – June 7, 2020, 15,380 individuals with pharmacy claims data and at least 6 months of enrollment were diagnosed and hospitalized with Covid-19 in UHG’s Clinical Discovery Database (Figure 1). Overall 52.8% were female, median age 70 years (IQR 58-80). Of these, 6,256 (40.7%) had a diagnosis of obesity or T2DM, of whom 1,185 (18.9%) died; and 2,333 (37.3%) had a metformin prescription. Individuals taking metformin were younger (73 vs 76 years), more often male (51.6%% versus 44.6%), and fewer died during their hospitalization for Covid-19 (17.8% vs 21.3%). Most persons taking metformin had T2DM (99.3%) and hypertension (56.3%). About 4% of persons taking metformin had asthma, 6% had chronic kidney disease, 19.5% had coronary artery disease, and 4% had liver disease. Individuals on metformin were less likely to have a history of venous thromboembolism (2.7% vs 4.1%), (Table 1). In unadjusted analyses, metformin was associated with decreased mortality, OR 0.802 (0.701, 0.917) (Table 2).

### Multivariate analyses of mortality by metformin use, in men and women with T2DM or obesity

Results of multivariate analyses are presented in Table 2. Mortality associated with all the variables in the multivariate analyses are shown in Tables S1-S3. The log-log plot assessing the proportionality assumption is presented in Figure s1; the distribution of the estimated propensity-score is in Figure s2; the standardized difference in propensity-matched covariates is in Figure s3. Metformin use not associated with statistically significantly decreased mortality in the overall sample (Table 2).

### Assessment for heterogeneity of effect by sex, in subgroups of women with T2DM or obesity

A Kaplan-Meier curve of survival by metformin use in men and women is shown in Figure 2. Metformin use was significantly associated with decreased mortality in women in logistic regression, OR 0.792 (0.640, 0.979); mixed effects model, OR 0.780 (0.631, 0.965); Cox proportional-hazards model, HR 0.785 (0.650, 0.951); and propensity-matched model, OR 0.759 (0.601, 0.960), (Table 2, Figure 3).

### Sensitivity analyses in patients with Covid-19 disease confirmed by polymerase chain reaction

In unadjusted analyses men and women with T2DM or obesity, metformin was not significantly associated with decreased mortality, OR 0.859 (0.737, 1.002), nor by the Cox proportional hazards shared frailty model in women, HR 0.808 (0.651, 1.003). Metformin was significantly associated with decreased mortality in women with type-2 diabetes or obesity in the minimally adjusted Cox shared frailty model, OR 0.790 (0.637, 0.978), and the propensity matched model, 0.744 (0.565, 0.980).

### Multivariate analysis of anti-TNFα inhibitors

Of the 15,362 persons included in the main analyses, 38 (0.25%) had claims for a TNFα inhibitor. In Cox proportional-hazards model, TNFα inhibitors were non-significantly associated with decreased mortality, HR 0.350 (0.087, 1.415). In a propensity matched model, matched for the same variables as the metformin analyses, TNFα inhibitors were non-significantly associated with decreased mortality, 0.483 (0.0821, 2.845). In a propensity model matched only on age, sex, charlson co-morbidity index, inflammatory bowel disease, rheumatoid arthritis and systemic lupus erythematosus, TNFα inhibitors were significantly associated with decreased mortality, OR 0.19 (0.038, 0.983), (Figures s4, eTable 4). A number of other variables were associated with increased or decreased risk of death from Covid-19, notably inflammatory bowel disease and asthma treated with beta2-agonists (Tables S1-S3).

## Discussion

This is the first study to report decreased mortality with outpatient metformin use in women with T2DM or obesity in a large cohort of patients hospitalized in the US for Covid-19, and to describe a sex difference in this response to metformin. These findings could have wide-reaching effects, as over 42% of women in the US have obesity.^29^ We found that metformin use was associated with significantly lower mortality among women across all multivariate analyses: logistic regression, mixed-effects analysis, Cox proportional-hazards, and propensity-matched models. The significant protective benefit in women compared to men may shed light on the mechanism by which metformin decreases mortality from Covid-19, as metformin has been shown to reduce TNFα in females more than males.^17-20^

We also found reduced mortality in persons with outpatient use of TNFα inhibitors who were hospitalized for Covid-19. TNFα inhibitor use was associated with large decreases in odds of mortality, but these findings were not statistically significant, likely because of the small sample size of 38. Reduced mortality in persons who use TNFα inhibitors would support previous research that TNFα plays a large role in the pathology of Covid-19.^30^ TNFα leads to macrophage activation and increased cytokine release, likely contributing to Covid-19 pathology.^31^

We considered other (overlapping) mechanisms by which metformin could reduce the severity of SARS-CoV-2 infection: ACE2 receptor modulation (via AMPK), decreased cytokine release (IL6, TNFα, increased IL-10), improved neutrophil to lymphocyte ratio, decreased glycemia (via AMPK), mast cell stabilization, decreased thrombosis, and improved endothelial function.^16-18,32-44^ In patients with and without diabetes, metformin has been shown to decrease inflammatory mediators IL-6 and TNFα.^16,43,45^ These effects are notable, as IL-6 and TNFα are thought to contribute to Covid-19 pathology.^13^ Metformin’s effects on these cytokines have been shown to differ by sex, with favorable effects in female over male mice, particularly for TNFα.^18^ Our findings of a strong sex-specific response to metformin in Covid-19 suggests that TNFα reduction may be the primary way by which metformin reduced mortality from Covid-19.

Our sex-specific findings are consistent with prior literature showing mortality benefit in women but not men from colorectal cancer.^20^ Possible reasons for sex-specific effects of metformin include the influence of sex hormones and epigenetic changes on the Y chromosome.^46^ There are 2 other potential ways in which metformin might cause sex-specific responses in Covid-19: Metformin inhibits IgE-and aryl hydrocarbon-mediated mast cell activation,^47^ and mast cell activation has been implicated as an early indicator of inflammatory response to SARS-CoV2 and possibly cytokine storm.^48^ Mast cells from female rats cause a greater increase in TNFα than mast cells in male rats, which could be one reason for greater benefit from metformin in women than men.^49^ Lastly, activation of AMPK by metformin can lead to increased expression of ACE2 and conformational changes to ACE2, and possibly decreasing SARS-CoV-2 binding to the ACE2 receptor.^35-37,50^ Recent work by Li et al found that expression of ACE2 receptor was equal in male and female human lungs, but that cytokine responses differed between men and women.^38^ This difference in subsequent inflammatory response and our sex-specific findings may support metformin’s anti-inflammatory effects as the primary means of benefit in Covid-19.

Additionally, in our multivariate analyses, beta2-agonist use was associated with decreased mortality in patients with asthma across all analyses (Tables S1-S3). This benefit may come from beta2-agonists’ effect on boosting IL-10, which is a predominantly anti-inflammatory cytokine that can reduce levels of TNFα.^51^ Metformin has also been shown to boost levels of IL-10, in females more than males.^43,45^ The mortality benefit in persons with asthma on beta2-agonists, combined with our finding of mortality benefit from metformin use, suggest that IL-10 may also be important in Covid-19.

In summary, we found that metformin was associated with a significant decrease in mortality for women with T2DM or obesity who were hospitalized for Covid-19, in an observational analysis of de-identified claims. We found no significant mortality benefit in men with T2DM or obesity who were hospitalized with Covid-19. The sex-specific effects of metformin on TNFα, IL-6, and IL-10, and our findings of benefit in women, might indicate that metformin’s protective effect in Covid-19 is primarily through effects on TNFα, IL-6, and IL-10. The importance of TNFα in Covid-19 is supported by our finding that TNFα inhibitor use was associated with decreased mortality from Covid-19. The fact that these outpatient medications convey benefit in patients hospitalized for Covid-19 is interesting because metformin is universally stopped at hospital admission, suggesting its protective effects begin prior to hospitalization.

Given metformin’s good safety profile and availability,^52^ it should be prospectively assessed for protective benefit from Covid-19. Gastrointestinal side effects from metformin can be eliminated for over 85% of patients with use of the newer extended-release formulations, and further with administration at the end of a meal.^53^ With the median time to hospitalization for Covid-19 being about 1 week, it is necessary to understand the duration of metformin use that conveys benefit, and whether it prevents Covid-19. Over 34% of adults in the US have prediabetes,^54^ yet it is highly under-diagnosed as over 90% of them are not aware that they do.^55^ Thus metformin for prevention of Covid-19 may be reasonable as metformin may be indicated for many adults in the US anyway. Additionally, increased inflammation from adiposity, and obesity’s association with increased risk of poor outcomes from Covid-19, metformin should also be assessed in patients of all BMI categories.

## Limitations

Our study has several limitations. Although claims data show metformin prescribed as a home medication for at least 90 days within the last 12 months, it does not give information about adherence. Metformin is sometimes purchased without insurance claims in this population because of its low cost, thus some individuals in our control group may have been exposed to the treatment, which would reduce the observed effect size. Prescriptions for outpatient use of metformin cannot be extrapolated to starting metformin at Covid-19 diagnosis or inpatient use. Retrospective analyses are subject to biases and unmeasured confounding. While multicollinearity among potential confounders makes interpreting their independent associations with in-hospital mortality potentially difficult, adjustments for these confounders is necessary to minimize bias for estimating the causal effect of home metformin use on in-hospital mortality.^56,57^

## Conclusion

In a large de-identified claims database of adults with T2DM or obesity, metformin was associated with significantly decreased mortality in women hospitalized with Covid-19, with no significant mortality reduction in men. Mechanistic reasons that support a sex-specific reason for metformin to be protective in Covid-19 include anti-inflammatory effects on TNFα, IL-6, and possibly IL-10. We also found that TNFα inhibitors were associated with reduced mortality, this finding was not significant in all models, likely due to the small sample size. Metformin has a good safety profile, availability, and needs to be prospectively assessed to understand mechanism, duration, and timing of treatment necessary for benefit. Given obesity’s pro-inflammatory effects that contribute to Covid-19 pathology, and the potential anti-inflammatory benefit of metformin in Covid-19, metformin should also be assessed in all BMI categories.

## Data Availability

The senior author has had access to the data for the entirety of the research process and analyses. The first author has had access to all of the data analyses.

---

**eFigure 1.**
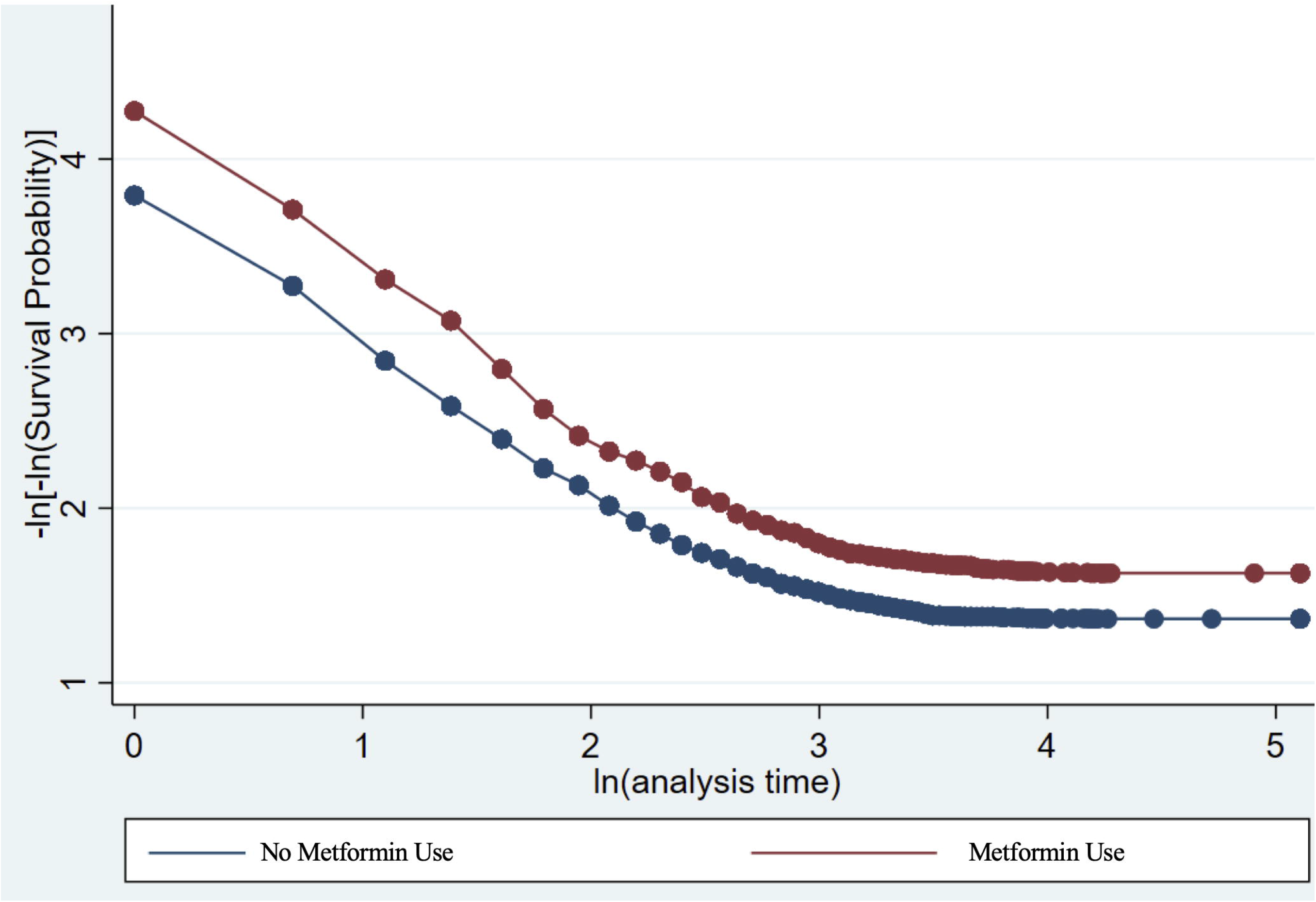
Log log plot assessing the proportionality assumption. A log-log plot assessing the proportionality assumption. The blue line represents no metformin use, the red line represents metformin use.

**eFigure 2.**
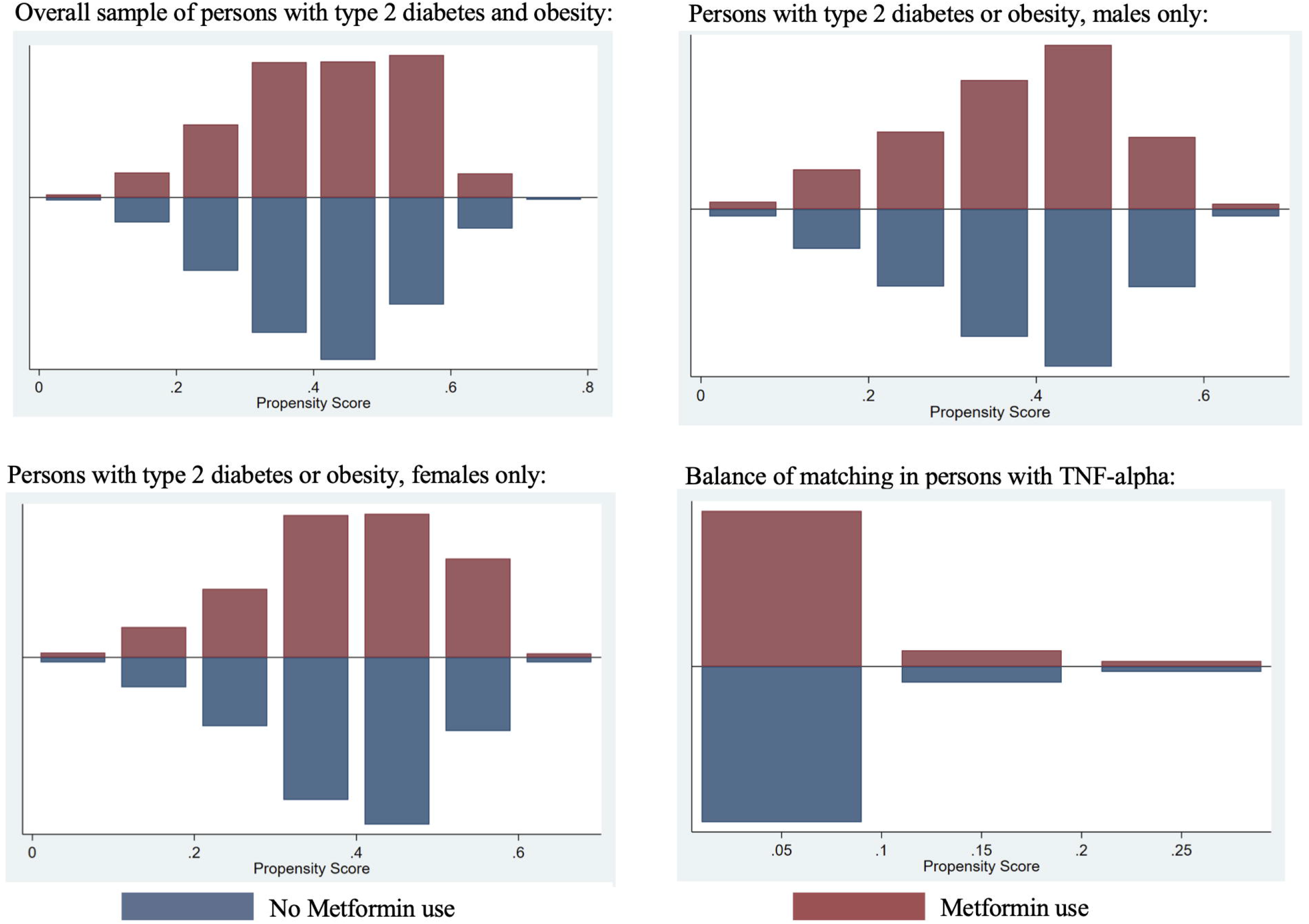
Balance of matching with caliper 0.2. A balance of matching of covariates in the propensity models. The blue represents no metformin use, the red line represents metformin use in all panels. The top left panel is the overall sample. The bottom left panel is women with type 2 diabetes or obesity. The bottom right panel is men with type 2 diabetes or obesity. The top right panel is among patients with home TNF-alpha inhibitor use.

**eFigure 3.**
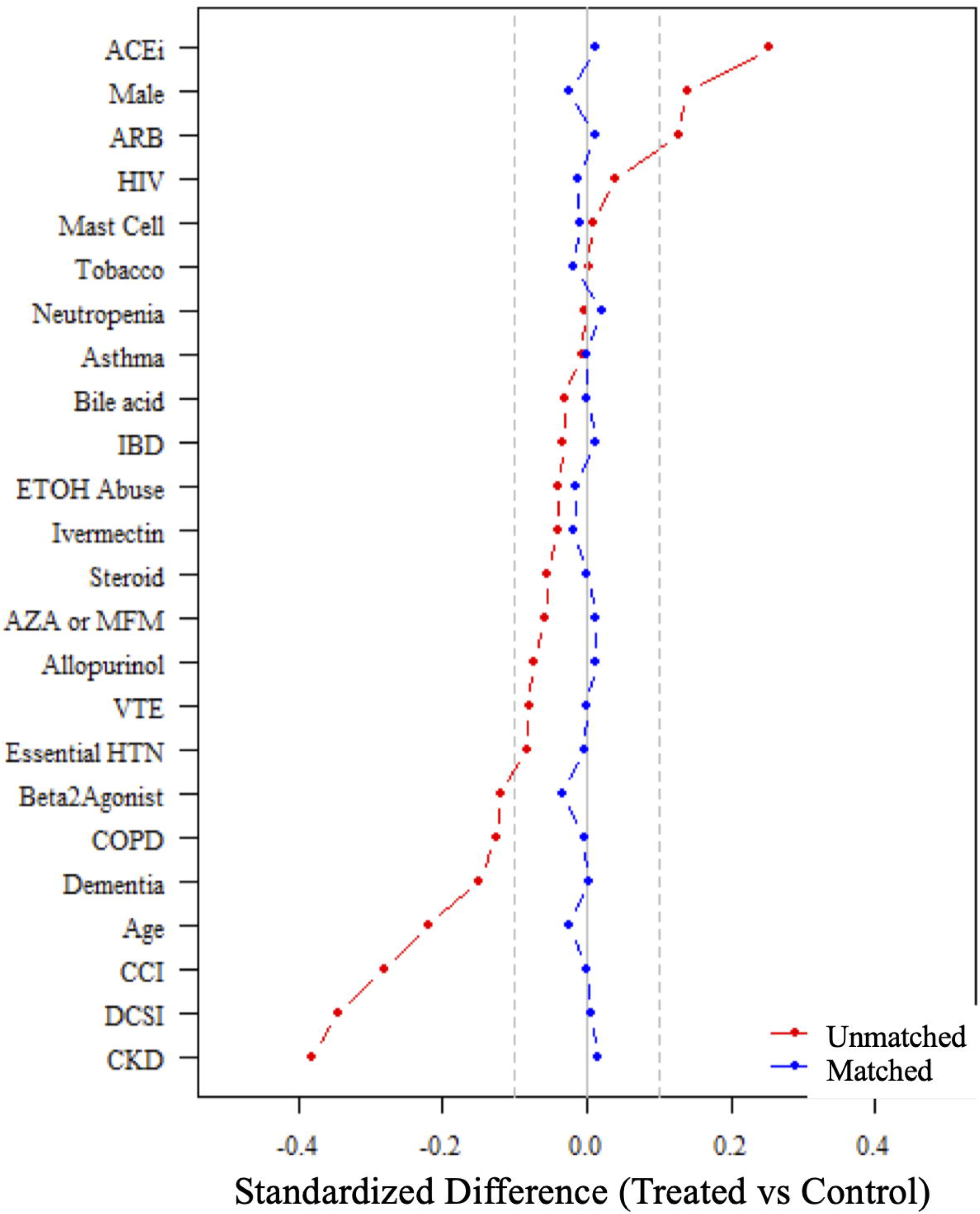
Propensity matched covariate balance. **Abbreviations:** ACEi=angiotensin converting enzyme inhibitor; ARB=angiotensin 2 receptor blocker; IBD=inflammatory bowel disease; ETOH=alcohol; AZA=azathioprine; MFM=mycophenolate mofetil; VTE=venous thromboembolism; HTN=hypertension; COPDc=chronic obstructive pulmonary disease; CCl=charlson comorbitidy index; DCSI=diabetes complications and severity index; CKD=chronic kidney disease. A forest line plot comparing standardized differences between covariates in the sample in the propensity matched models. The red line represents before matching and the blue line presents after matching.

**eFigure 4.**
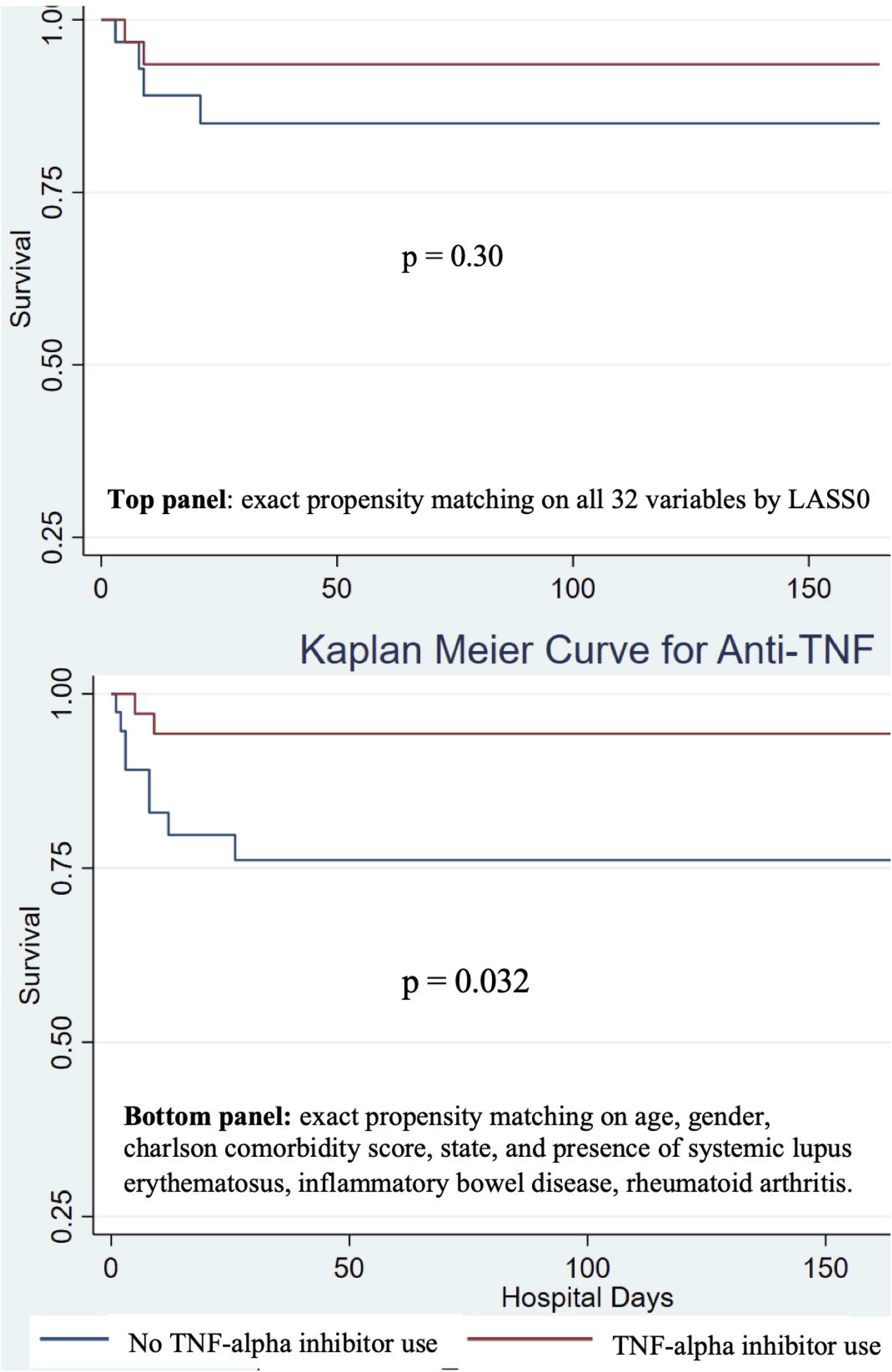
Kaplan Meier curve by 1NF-alpha inhibitor use, with propensity matching in persons hospitalized for Covid-19. Kaplan Meier survival curve comparing patients on home TNF-alpha inhibitors to those not on TNF-alpha inhibitors. The top panel shows survival when matched on all variables identified by LASSO. The bottom panel shows survival when matching on variables clinically relevant to TNF-alpha inhibitor use (age, gender, charlson comorbidity score, state, presence of systemic lupus erythematosus, inflammatory bowel disease, rheumatoid arthritis. The y axis is probability of survival, the × axis is number of days in the hospital. The blue line represents no TNF-alpha inhibitor use, the red line represents TNF-alpha inhibitor use.

